# Machine Learning Approaches to Determine Feature Importance for Predicting Infant Autopsy Outcome

**DOI:** 10.1101/2020.05.21.20105221

**Authors:** J Booth, B Margetts, W Bryant, R Issitt, C Hutchinson, N Martin, NJ Sebire

## Abstract

**Introduction:** Sudden unexpected death in infancy (SUDI) represents the commonest presentation of postneonatal death, yet despite full postmortem examination (autopsy), the cause of death is only determined in around 45% of cases, the majority remaining unexplained. In order to aid counselling and understand how to improve the investigation, we explored whether machine learning could be used to derive data driven insights for prediction of infant autopsy outcome.

**Methods:** A paediatric autopsy database containing >7,000 cases in total with >300 variables per case, was analysed with cases categorised both by stage of examination (external, internal and internal with histology), and autopsy outcome classified as ‘explained (medical cause of death identified)’ or ‘unexplained’. For the purposes of this study only cases from infant and child deaths aged <= 2 years were included (N=3100). Following this, decision tree, random forest, and gradient boosting models were iteratively trained and evaluated for each stage of the post-mortem examination and compared using predictive accuracy metrics.

**Results:** Data from 3,100 infant and young child autopsies were included. The naïve decision tree model using initial external examination data had a predictive performance of 68% for determining whether a medical cause of death could be identified. Model performance increased when internal examination data was included and a core set of data items were identified using model feature importance as key variables for determining autopsy outcome. The most effective model was the XG Boost, with overall predictive performance of 80%, demonstrating age at death, and cardiovascular or respiratory histological findings as the most important variables associated with determining cause of death.

**Conclusion:** This study demonstrates the feasibility of using machine learning models to objectively determine component importance of complex medical procedures, in this case infant autopsy, to inform clinical practice. It further highlights the value of collecting routine clinical procedural data according to defined standards. This approach can be applied to a wide range of clinical and operational healthcare scenarios providing objective, evidence-based information for uses such counselling, decision making and policy development.

## Introduction

Great Ormond Street Hospital for Children NHS Trust (GOSH) is the largest specialist centre for treatment and investigation of children in the United Kingdom, with UCL Great Ormond Street Institute of Child Health, representing the largest centre for paediatric research outside the United States. At GOSH specialist Paediatric Pathologists perform perinatal, infant and childhood postmortem examinations (autopsies/PMs), including hospital referrals, forensic cases and those on behalf of Her Majesty’s Coroner, including those for sudden unexpected death in infancy and childhood (SUDI/C), the commonest presentation of post-neonatal early childhood death. However, in this group despite comprehensive autopsy investigation, only around 45% of cases result in an identifiable medical cause of death, the majority remaining unexplained.[1],[2],[3]

The GOSH Pathology Department has established a research database containing structured details of all autopsies performed between 1996 and 2018. The database was originally developed for research into SUDI but has since been utilised for a number of other projects investigating stillbirth and various aspects of paediatric autopsy procedure.[4], [5] Currently the database holds data for >7,000 fetal and paediatric autopsies, with more than 300 data items defined for each postmortem examination. The data items record the four main stages of the autopsy; external examination, dissection and internal examination, then grouped by bodily system examined at both the macroscopic and microscopic histological level.

The database allows controlled use of deidentified information and it’s use for research has been approved by the appropriate Research Ethics Committee / IRB (REC approval 16LO1910, London, Bloomsbury).

The purpose of this project was to investigate feasibility of a data science approach, using routinely collected and deidentified autopsy data, to determine which elements are most contributory to determining a medical cause of death, in order to both develop future operational strategies to increase procedural efficiency and to provide objective information which could potentially be used both for planning and counselling parents and families. This included specifically; to extract data from the existing research database (MS Access) into an entity attribute value schema to optimise data analytics[6] and the efficiency of storing data[7] as well as flexibility for health care data, to apply a Decision Tree analytical method to the extracted data (Decision tree methodology is a commonly used data mining method for establishing classification systems based on multiple covariates or for developing prediction algorithms for a target variable).[8] We further explore ensemble methods which combine techniques to balance variance versus bias,[9] including Random Forests, in which training data is split into a number of different sets and a tree is calculated for each set and the results combined, and Gradient boosting, in which parameters that give a low prediction accuracy are combined to produce a higher prediction accuracy.[10]

## Methods

Data engineering was undertaken using the Python programing language,[11],[12] Initial data manipulation used structured query language (SQL) instigated using PyODBC,[13] which allows connection to databases using ODBC connections and the production and return of SQL queries. The initial step included creating concepts, events and attributes in the EAV schema and importing data from the MS Access research database. Each autopsy was regarded as a single event, with fields represented as event attributes. Summary event attributes were then calculated for reporting and analytic purposes added to the existing set of events. This allowed generation of four research data views (RDVs), one for each autopsy stage, in the form of CSV files. In order to optimise the data for machine learning we used One-hot encoding for categorical features (rather than each feature having a single column of data with the appropriate category; each category has its own column with either a 1 or 0 depending on whether each event has that feature value), and normalisation of numeric values such that each numeric value was normalised based on their predicted value for the age of the patient described by each event such that each numeric value will be in the range 0 – 1 with only outliers having larger values. This allowed production of the four final adjusted RDVs, one for each autopsy stage.

Analysis was undertaken using the R programming language[14].[15] In short, models were created using default parameters, (Appendix),which were then changed individually to obtain an optimal value based on predictive accuracy, and repeated to finalise a set of parameters for each autopsy stage for each model. The output for each of the modelling stages was an R function that can be called for that model with a training/test split which saves the resulting confusion matrices and relative feature importance, with plots carried out using ggplot2.[16] Rpart package was used for recursive partitioning on trees, for classification and regression to achieve an optimum level of complexity for a given set of data,[17] with subsequent Xtreme Gradient Boosting, an efficient implementation of the gradient boosting framework.[18] Using these functions, models were run for each model package for each autopsy stage for five different random seeds each deriving a separate training/test data split, which were combined to produce a comparison of model predictive accuracy for changing random seeds, comparison of change in predictive accuracy of each stage, comparison of relative feature importance changes for different random seeds for each stage, and a final predictive accuracy for determining a cause of death at each stage for a final set of relative feature importance by model by stage.

The results are assessed in terms of the predictive accuracy of the different models and then the relative feature importance and their change as the different stages of the postmortem examination.

## Results

Data from 3,100 autopsies were included in the analysis. The number of missing values by core variables and an example decision tree output are shown in Figure 1. Confusion matrices for decision tree, random forest and XG Boost models are shown in Figure 2. The overall performance to predict whether cause of death could be determined (combined) was greatest when data from all four stages was included and the XG Boost provided the best performing model, correctly classifying 80% of cases. (Figure 3) Both ensemble methods outperformed the basic decision tree model and the XGBoost model outperformed Random Forest but only by a small margin. The underlying increase on predictive accuracy as the post-mortem stages progress is reflected across all three models. Since XGBoost provides the best classification performance, determination of feature importance is presented from this model (Figure 4).

**Figure 1.**
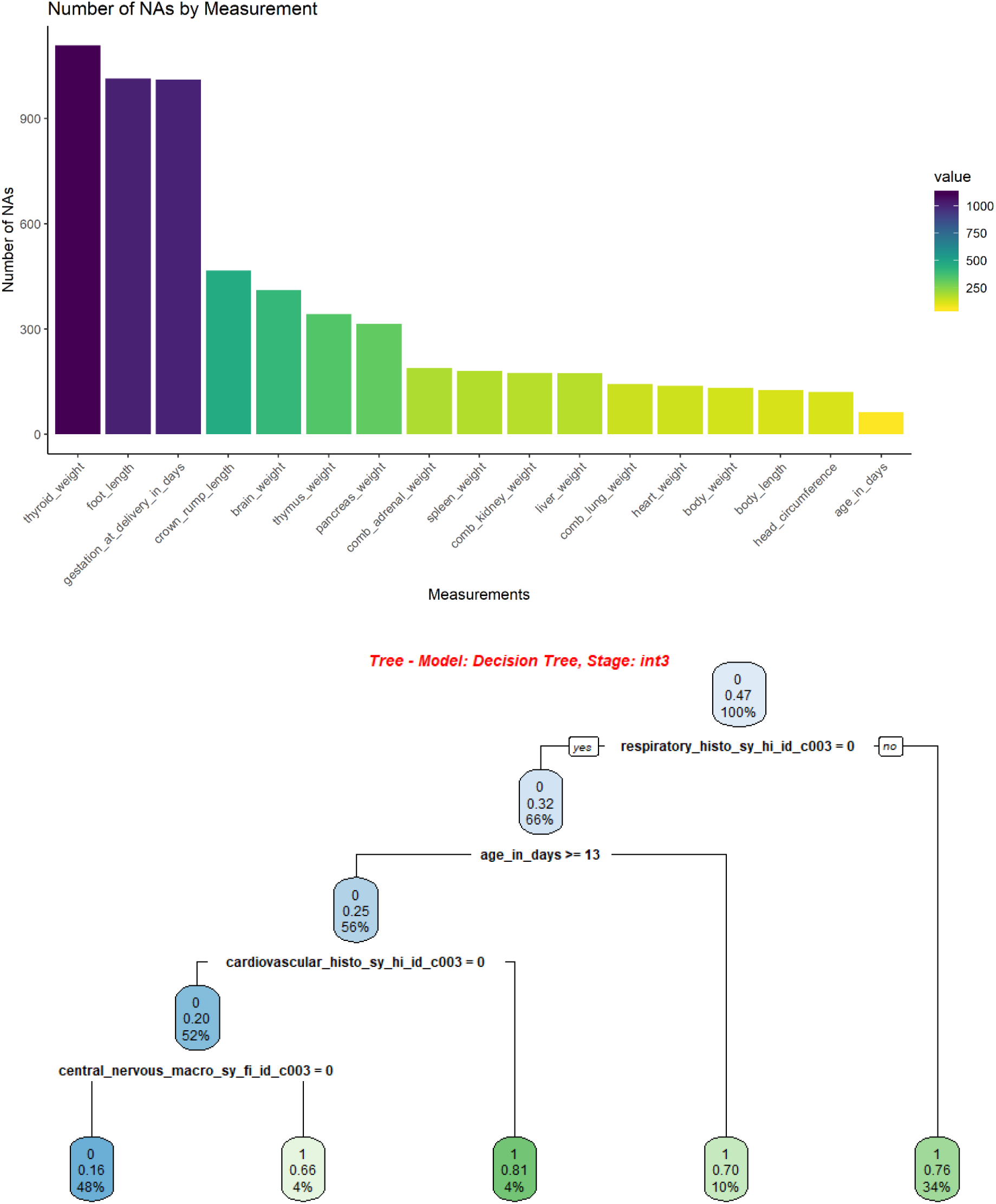
Chart illustrating numbers of missing values for main variables during initial data preparation (Top), and example initial decision tree output diagram (Bottom)

**Figure 2.**
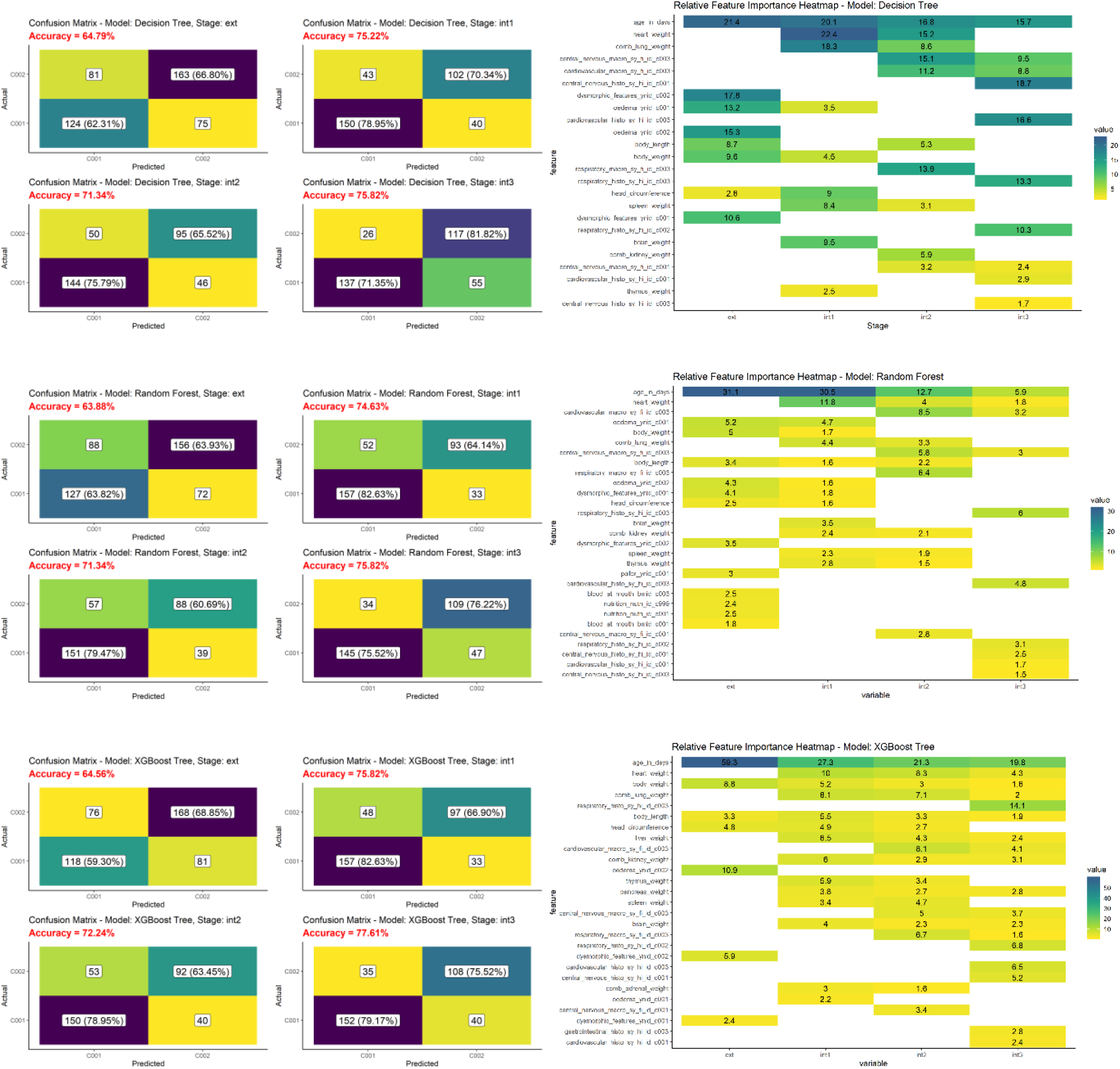
Confusion matrices (Left) and relative feature importance plots (Right) for decision tree (top), random forest (middle) and XGBoost (bottom) models.

**Figure 3.**
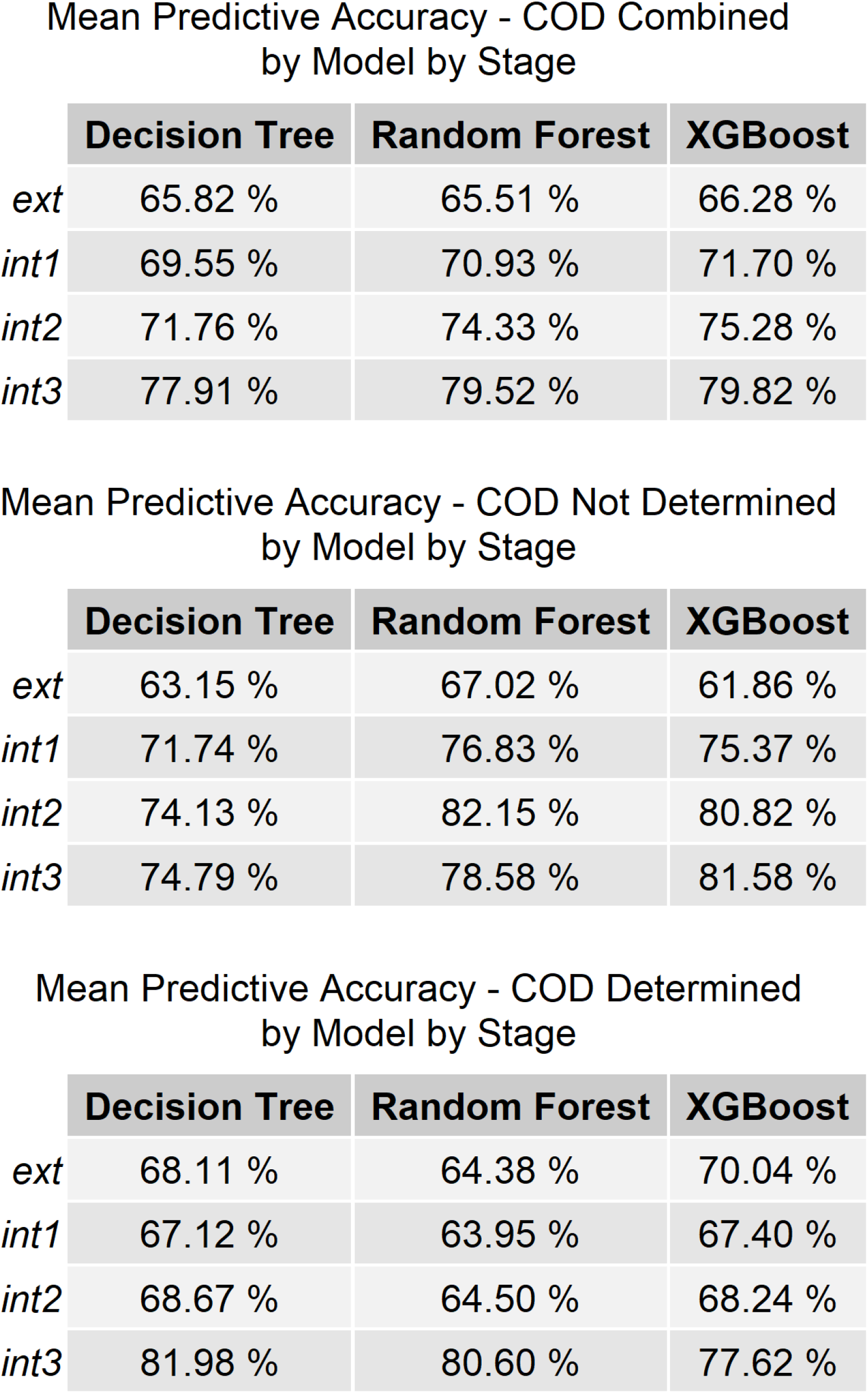
Mean predictive accuracy tables for three models demonstrating overall best performance for the XGBoost model using all stages of the autopsy.

**Figure 4.**
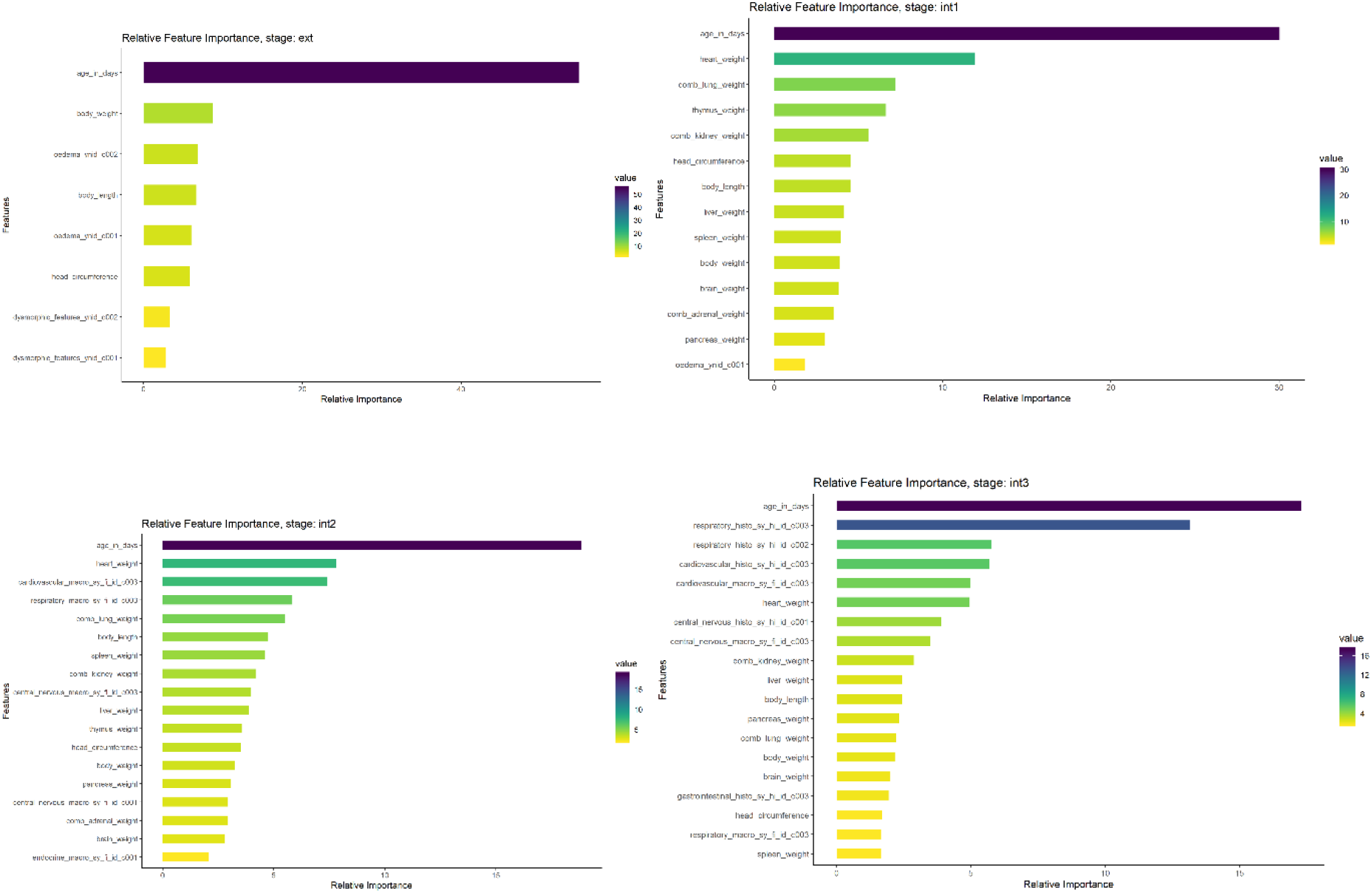
Relative feature importance of the final XGBoost model at all stages, demonstrating the main influence of age and respiratory/cardiovascular histology for likelihood of determining cause of death.(Ext=external (top left), int1 (biometry; top right), 2 (macroscopic findings; bottom left), 3 (histological findings; bottom right) = first, second and third stages of internal examination respectively)

At the initial (external examination) stage of the postmortem examination, only the age of the patient has significant bearing on being able to determine the cause of death, with decision tree output determining the main boundary as around 16 days of age, with a secondary boundary of around 276 days. At the second stage, initial internal examination, age remains of primary importance but organ weights begin to have significance, decision tree output suggesting the feature boundary being variation from ‘normal’ of >30%. However, importantly, once histological findings are available, in the final stage, these histology classifications now play the most important role after age, especially histological findings of respiratory and cardiovascular systems.

## Discussion

The findings of this study have demonstrated that it is possible to evaluate a clinically large structured dataset derived from routine clinical data, using machine learning methods in order to identify key components of a medical investigation procedure which provide most value, in this case infant autopsy, in relation to predicting whether a medical cause of death is determined. The advantage of such an approach is the objective determination of feature importance based on findings from the data set, with less dependence on medical practitioner opinions or presuppositions (although of course these are inherent in some feature identification), and this therefore represents a potentially powerful approach for future evaluation of care pathways and complex procedures. The key advantage of the Decision Tree technique is it simplifies complex relationships between input variables and target variables by dividing original input variables into significant subgroups, thus making the model easier to understand and interpret. The main disadvantage of the technique is that using a single tree a model will suffer from low variance and high bias.[19]

The machine learning approach identified age as an important factor predicting the likelihood of determining a medical cause of death, specifically, cases with age<16 days or more than 276 days at death being more likely to be associated with a medical cause identified. Interestingly, this is in agreement with the previous observations that cases of sudden unexpected early neonatal death in the first 7 days of life, represent a distinct group of infant deaths with a greater likelihood of being explained and including different causes of death such as inherited metabolic disease,[20], [21] and that cases of sudden unexpected death in childhood, in children over one year of age, are also more likely to be associated with a medical cause of death being identified.[22], [23] However, whilst previous age cut-off boundaries were purely empirical (7 days and 365 days), the current data suggest that, whilst broadly similar, the boundaries may be more appropriate at 16 days and 276 days of life.

Initial internal examination and dissection was of some importance, particularly whether organs are enlarged, such as increased heart weight, which is in accordance with previously reported findings in autopsy cases of myocarditis.[1] However, once available, histological findings, in particular of respiratory and cardiovascular systems play important roles. These findings are in agreement with those reported from previous autopsy studies which suggest that histological examination of heart and lungs provide by far the most important information for determining cause of death with very little value of routine histological examination of the majority of other organs.[24], [25],[26]

Whilst the results presented here are those from the machine learning process and classification model, this study has also highlighted the importance of data engineering required to prepare routine clinical data sets for machine learning use, the importance and complexity of this task often being underestimated by researchers.[27], [28] Specifically, determining a data model and developing the initial extract, transform and load (ETL) process and preparation of the adjusted RDV structures to be suitable for use by all three of the modelling packages, particularly age normalising measurement features as part of the project pipeline process. Furthermore, the creation and tuning of three model types for four stages of the autopsy represents significant investment; developing a well-tuned model on real world data is a non-trivial task.

The advantages of this study are the large number of cases included with an extensive, well characterised and unified clinical data, representing a unique population resource, in addition to a systematised and well described approach to data engineering and machine learning evaluation. The main limitations relate to the use of real world data; in other words, data was collected at the time of routine autopsy examination and whilst objective criteria were used to categorise findings, many features such as whether histological examination was normal or abnormal, were dependent on the interpretation of the pathologist undertaking the autopsy at the time. In addition, it should be noted that for the purposes of this study the features were identified in order to classify whether or not the final cause of death would be explained or unexplained, but not to determine any of the specific causes of death within the explained group. Additional analysis, using an extended dataset with additional ancillary investigations could allow prediction of specific causes of death rather than binary determination of explained versus unexplained. Finally, through use of unsupervised multidimensional clustering approaches, such as tSNE, it may be possible to identify distinct sub groups within a larger population such as those cases in which the cause of death remains unexplained, which could lead to new hypotheses for future investigation.[29], [30]

In summary, this study has demonstrated the use of a machine learning model related to classification of determination of cause of death following autopsy in infants and young children, with an XGBoost decision tree model providing best performance. Through this model, the main objective factors for predicting whether a cause of death will be determined at each stage of the autopsy have been identified, with the most informative factors being age less than 16 days or more than 276 days at death, and abnormal cardiovascular or respiratory histological features. The majority of other investigations at autopsy provide little additional information. The findings provide an objective evidence based approach for determining policy and counselling of families, and it is highly likely that similar machine learning approaches can be used in a range of complex medical investigation settings.

## Data Availability

Nonidentifiable aggregate data can be obtained from the authors

## Declarations

### Compliance with Ethical Standards

The work complies with all ethical standards and was approved by the Bloomsbury (London REC)

### Author contributions

NJS, JCH and JB were involved in autopsy data collection and database establishment and development. NJS and JB conceived the project. JB, BM, RI, WB and NM were involved in running the data analysis and generating results. All authors were involved in writing and preparing the manuscript.

## Appendix

Parameters of the R functions were:

Decision Tree:

- minsplit - The minimum number of observations that must exist in a node in order for a split to be attempted.
- minbucket - The minimum number of observations in any terminal node. Use minsplit / 3.
- cp – Complexity parameter, used to define further pruning after the initial tree is produced.

Random Forest:

- Mtry - Number of candidates draw to feed the algorithm. By default, it is the square of the number of columns.
- Maxnodes - Set the maximum amount of terminal nodes in the forest.
- ntree - Number of trees in the forest.

XGBoost:

- Eta – Controls how much information from a new tree is used in boosting.
- max_depth – Controls the maximum depth of a tree.
- gamma - Controls the minimum reduction in the loss function required to grow a new node in a tree.
- min_child_weight - Controls the minimum number of observations (instances) in a terminal node.
- Subsample - This parameter determines if we are estimating a Boosting or a Stochastic Boosting.
- colsample_bytree - Number of features to sample in each new tree.

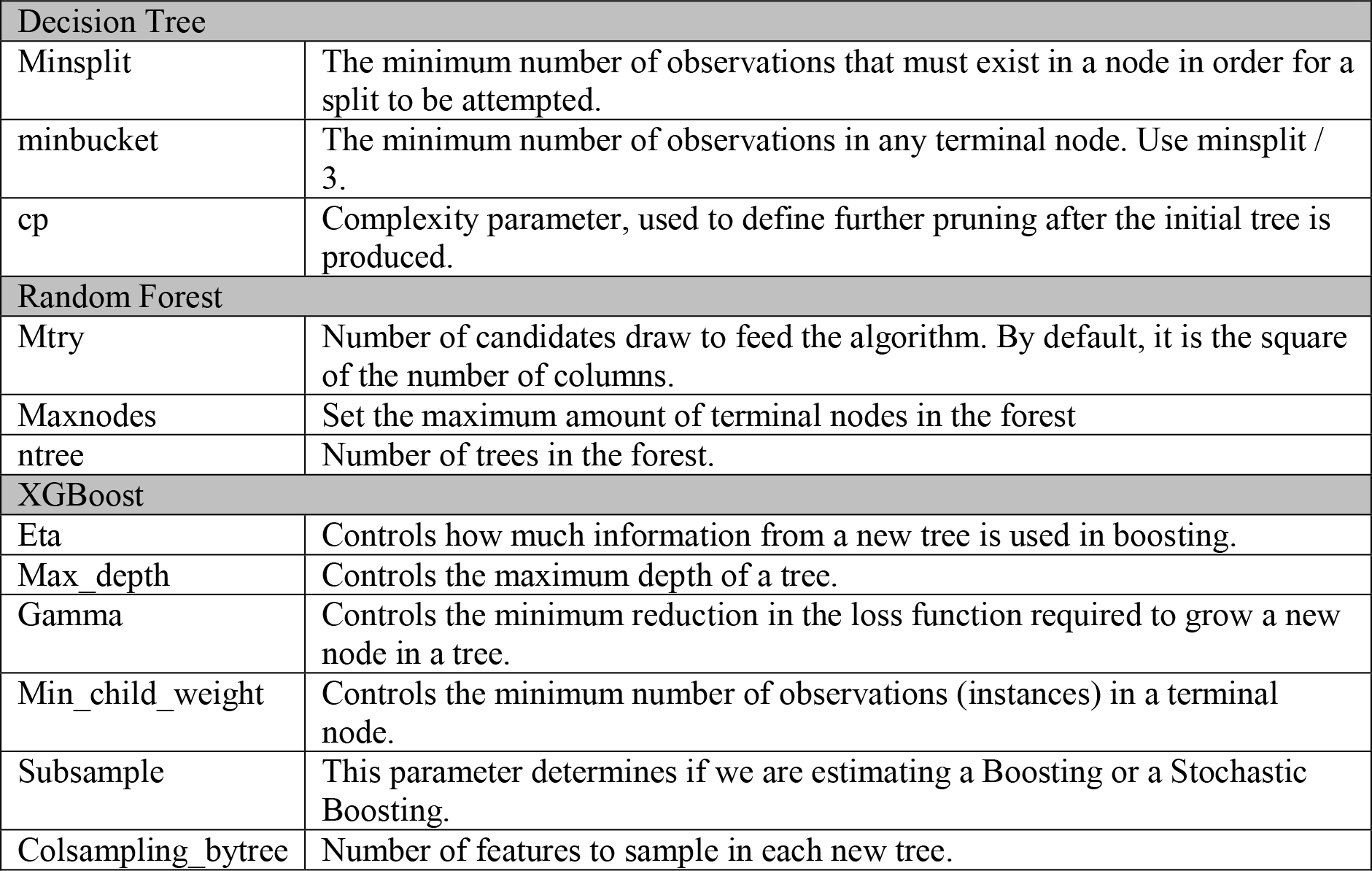

